# Optimizing high-intensity interval training volume in heart failure: dose–response meta-analysis of randomized controlled trials

**DOI:** 10.1101/2025.06.02.25328596

**Authors:** Luis Suso Martí, Miriam Garrigós-Pedrón, Juan J. Amer-Cuenca, JF. Lisón, Carlos Salvador Huerta, Óscar Fabregat-Andrés, Daniel López-Fernández, Joaquín Calatayud, Noemí Valtueña-Gimeno, Francisco José Ferrer-Sargues

## Abstract

**Background:** Chronic heart failure (CHF) is a leading cause of morbidity and mortality worldwide, associated with impaired functional capacity and quality of life. Although exercise-based cardiac rehabilitation programs, especially high-intensity interval training (HIIT), have been shown to be beneficial, the optimal duration of HIIT remains unclear. This study aimed to determine the HIIT volume that maximizes VO_2_max improvement in CHF patients, in terms of both session and total duration.

**Methods:** A systematic review and dose–response meta-analysis of randomized controlled trials was conducted to assess the effects of HIIT on maximum oxygen consumption (VO_2_max) in CHF patients. Articles selected measured VO_2_max pre and post HIIT intervention with a cardiopulmonary exercise test. Articles including patients who had undergone surgery for pacemaker or ventricular assist device implantation were excluded. The risk of bias of the studies was assessed using the RoB 2 tool. Restricted cubic spline models were used to explore dose–response relationships for both total exercise minutes and session duration.

**Results:** Twenty-one studies comprising 449 participants were included. HIIT significantly improved VO_2_max (mean difference: 3.19 mL/kg/min; 95% CI: 2.29–4.09). A non-linear dose–response relationship was identified. A minimum dose of 30 minutes per session and 900 total minutes was necessary to achieve a minimal clinically important difference. The greatest improvements in VO_2_max were observed with 50-minute sessions and a total of 1.300 exercise minutes.

**Conclusions:** HIIT is effective in improving VO_2_max in CHF patients. However, the optimal improvement of cardiorespiratory fitness in this population is directly related to the time and frequency of the training. Standardizing and adjusting the volume of the programs can ensure effective rehabilitation and improve patient adherence. These results provide evidence-based practical guidance for clinicians to design more efficient and targeted interventions for this high-risk population.

**CRediT authorship contribution statement:**

- Conceptualization, methodology, formal analysis, investigation and writing-original draft and final approval of the version to be published: Luis Suso Martí; Francisco José Ferrer-Sargues; Noemí Valtueña-Gimeno.
- Methodology and Writing-review and editing: Miriam Garrigós-Pedrón; Juan J. Amer-Cuenca; JF Lisón; Joaquín Calatayud.
- Investigation, Visualization and Writing-review and editing: Carlos Salvador Huerta; Óscar Fabregat-Andrés; Daniel López-Fernández.

## INTRODUCTION

Chronic heart failure (CHF) represents one of the leading causes of morbidity and mortality worldwide, affecting millions of individuals.^1^Projections show that the prevalence of CHF will increase 46% from 2012 to 2030, with an estimated prevalence of 3% of the population by 2030.^2^ Characterized by left ventricular systolic and/or diastolic dysfunction, CHF is defined not only by cardiovascular impairments but also by significant peripheral abnormalities, including skeletal muscle dysfunction (e.g., sarcopenia) and endothelial impairment.^3^ However, among these features, cardiorespiratory fitness is also one of the most important independent predictor of CHF prognosis and quality of life.^4^Directly measured by maximum oxygen consumption (VO_2_max), this measure has been related as a strong prognostic marker for better prognosis and higher survival rate in CHF patients.^5^ In fact, it has been suggested that it is more cardioprotective than the overall physical activity levels. ^6^

In this context, exercise-based cardiac rehabilitation programs (CRP) are considered a fundamental therapeutic approach in the management of CHF.^7^ Randomized clinical trials and meta-analyses have demonstrated that exercise, particularly aerobic training, improves functional capacity, enhances quality of life, and reduces hospitalizations in patients with CHF.^8,9^ However, in recent years, high-intensity interval training (HIIT) has emerged as a promising therapeutic strategy for cardiovascular patients, demonstrating efficacy and safety as a core component of CRP. ^10,11^ Its adaptability—spanning supervised hospital-based protocols to home-based interventions—further underscores its clinical utility in improving cardiovascular outcomes. ^10,11^

For instance, HIIT has been shown statistically significant and clinically relevant effects on improving VO_2_max, either alone or in combination with other training modalities.^12^ This training modality has shown better effects compared to moderate-intensity continuous training in terms of VO_2_max and in cardiovascular and peripheral adaptations.^13^ In addition, different studies have proved that many individuals report equal or greater enjoyment with HIIT and showed at least similar overall training adherence compared with moderate-intensity continuous training and it is also relatively time-efficient.^14^

But to quantify the beneficial effects of HIIT, and to optimize the exercise prescription in a reliable and reproducible manner, it is fundamental to standardize the prescription parameters through a global consensus. The American College of Sports Medicine (ACSM) has proposed a model for prescribing exercise based on four main parameters (frequency, intensity, time and type of exercise). This model has been named FITT principle, and has been adapted to different populations, both healthy subjects and those with different chronic illnesses.^15^ACSM and the American Heart Association, among others, have stablished and adapted this FITT principle to cardiac populations, such as those suffering CHF. Among these prescriptive parameters, the total exercise volume of CRP represents a critical determinant for eliciting optimal physiological and clinical adaptations. However, to our knowledge, no dose-response meta-analysis has been performed regarding optimal HIIT volume in CHF.

Thus, the present meta-analysis aims to determine the optimal HIIT volume for maximizing VO_2_max in patients with CHF, in terms of both session and overall program duration. To the best of our knowledge, this is the first dose-response study of HIIT in this population, and the objective is to provide robust evidence to optimize the CRP prescription parameters in this high-risk group.

## METHODS

### DESIGN

This systematic review and dose-response meta-analysis was performed under the recommendations of Preferred Reporting Items for Systematic Reviews and Meta-Analysis (PRISMA).^16^ The systematic review protocol was registered in the International Prospective Register of Systematic Reviews (PROSPERO) with the number (BLINDED).

### SEARCH STRATEGY

A systematic literature search of PubMed/MEDLINE, EBSCO and Google Scholar was performed independently by two reviewers. The group of keywords combined for searching were “heart failure”, “high intensity interval training”, “maximum oxygen consumption” and “peak oxygen uptake”. Only Randomized Controlled Trials were included. The last search was conducted on July 22, 2024. The articles retrieved were exported to the systematic review manager Rayyan (rayyan.ai) for further analysis.

### ELEGIBILITY CRITERIA

This systematic review includes studies on the effects of HIIT on exertion performance in CHF patients. Eligible criteria were studies that included adult patients (aged ≥ 18 years) diagnosed with heart failure and randomized for a HIIT intervention. The outcome of interest was VO_2_max measured with CPET. Pre-intervention measurement was used as a comparator with VO_2_max after intervention to quantify the value improvement. No restriction to the intervention time was defined as inclusion criteria. Studies that included patients who had undergone surgery for pacemaker or ventricular assist device implantation were excluded. Also sub studies of SMARTEX Study and OptimEx-Clin Study, already included in this revision, were excluded.

### SELECTION CRITERIA

Following the removal of duplicates using the system review manager, two independent reviewers were tasked with screening the table, abstract and keywords for each study. Then, full text review of the remaining articles was conducted to ensure they met the inclusion criteria. In the event of a discrepancy among reviewers, a third reviewer was assigned to arbitrate.

### DATA EXTRACTION

The data was extracted using standard data extraction forms of Cochrane guidelines.^17^ The following variables were considered: first author, year of publication, number of participants and pre- and post-intervention VO_2_max means and standard deviations (SD). To determine the specific HIIT dose, total exercise time was extracted, disregarding the warm-up and cool-down minutes. If exercise time per session was not reported directly in the article, it was calculated by the details of the training protocol.

### RISK OF BIAS ASSESSMENT

The risk of bias assessment for each study included in this meta-analysis was conducted independently by two reviewers. The Cochrane risk-of-bias tool for randomised trials, version 2 (RoB-2)., was used in this process.^18^ The domains assessed for the risk of bias were randomization process (D1), deviations from intended interventions (D2), missing outcome data (D3), measurements of the outcome (D4) and selection of the reported result (D5). Finally, an overall score was set for each RCT. For each domain, a judgment was assigned with a “Low” answer to indicate a minimal risk of bias, a “High” indicating an elevated risk of bias, or “Some Concerns” indicating an unclear or unknown risk of bias where studies had reported insufficient details.

### DATA SYNTHESIS AND ANALYSIS

All analyses were performed with RStudio software (RStudio, PBC, Boston, MA). First, the effect of HIIT on VO_2_max was analysed in a random-effects pre-post meta-analysis. Mean differences (MDs) and their corresponding 95% confidence intervals (CIs) were obtained. Heterogeneity was assessed using the I² statistic. Heterogeneity of 0-40% was defined as unimportant, 30-60% as moderate heterogeneity, 50-90% as substantial heterogeneity, and 75-100% as substantial heterogeneity.^19^ To detect publication bias, a visual inspection of the DOI plots was performed to look for any asymmetry. Additionally, the Luis Furuya-Kanamori (LFK) index was used as a quantitative measure. An LFK index within ±1 represents no asymmetry, an LFK index above ±1 but within ±2 represents a small asymmetry, and an LFK index above ±2 represents a large asymmetry.^20^

Second, a one-stage dose-response meta-analysis was conducted to evaluate the dose-response associations between total minutes of HIIT (dose) and changes in VO_2_max (response).^20^ Thus, selected effect sizes and corresponding covariances (SDs) were used to estimate study-specific dose-response curves, using the MD of the within-group change from baseline to post-treatment measurement for VO_2_max and the total minutes of HIIT as the dose. The dose-response association was defined using a restricted cubic spline model with three nodes at the 10th, 50th, and 90th percentiles. The relative efficacy of the dose studied compared to a “zero” dose is characterized by dose-response curves.

## RESULTS

### STUDY SELECTION

The initial electronic search yielded 515 potentially eligible articles. After removing 280 duplicates, 180 studies were excluded following title and abstract screening. Subsequently, 69 full-text articles were assessed, and after applying the eligibility criteria, 21 RCTs were ultimately included. The study selection process is detailed in Figure 1.

**Figure 1.**
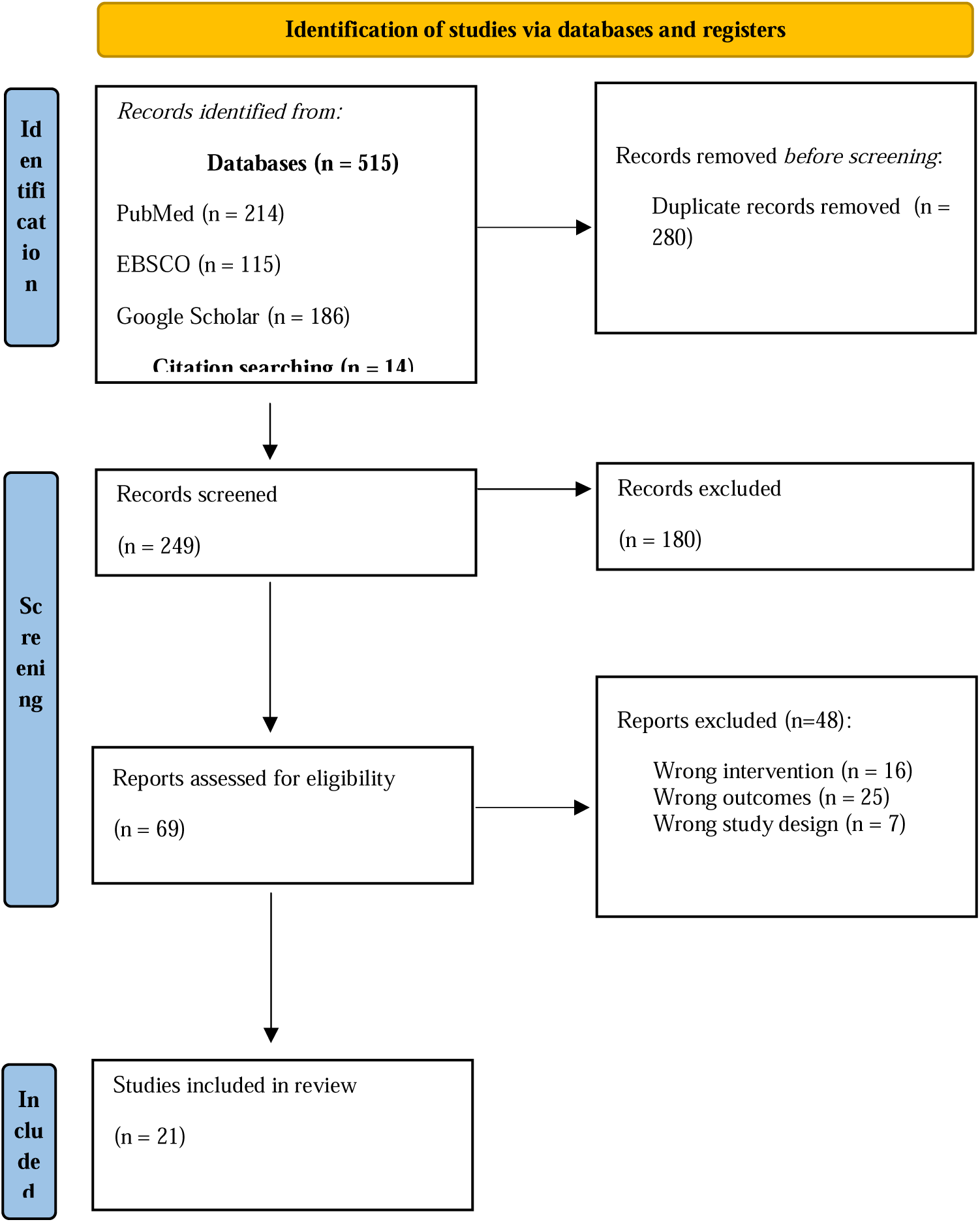
Preferred Reporting Items for Systematic Reviews and Meta-Analyses (PRISMA) flow diagram.

### CHARACTERISTICS OF THE STUDIES

Of the 21 trials,^21–41^ 449 patients (68% male) followed the HIIT based cardiac rehabilitation programs presented in the selected studies. Patients mean age was 60.5 ± 1.6 years and the mean left ventricular ejection fraction (LVEF) in % was 38.8 ± 5.3. In the 13 studies where the New York Heart Association (NYHA) functional class of heart failure ranging was reported, all the patients were between class I and III, not recording any IV class patient.^21,23,24,26–29,32,34,36–39^ The demographic and clinical characteristics reported by the study for each patient’s sample are shown in Table 1.

**Table 1.** Demographic and clinical characteristics of the patients reported in each study.

| Study | N Gender<br>(male/female) | Mean Age | Diagnosis<br>(NYHA)* | LVEF<sup>†</sup><br>(%) |
| --- | --- | --- | --- | --- |
| 1 Anagnostakou et al. | 14 M/F: 11/3 | $53 \pm 10$ | I/II/III: 5/7/2 | $39 \pm 11$ |
| 2 Angadi et al. | 9 M/F: 8/1 | $69 \pm 6.1$ | NR<sup>†</sup> | $63.7 \pm 6.4$ |
| 3 Benda et al. | 10 M/F: 9/1 | $63 \pm 8$ | II/III: 8/2 | $37 \pm 6$ |
| 4 Besnier et al. | 16 M/F: 11/5 | $59 \pm 13$ | I/II/III: 8/7/1 | $36 \pm 8$ |
| 5 Chou et al. | 17 M/F: 12/5 | $60.9 \pm 5$ | NR<sup>†</sup> | $36.1 \pm 5.2$ |
| 6 Chrysohoou et al. | 39 M/F:<br>28/11 | $56 \pm 11$ | I/II/III:<br>15/20/4 | $32 \pm 0.0$ |
| 7 Dimopoulos et al. | 10 M/F: 9/1 | $59.2 \pm 12.2$ | I/II/III: 3/6/1 | $34.5 \pm 10.5$ |
| 8 Donelli et al. | 10 M/F: 3/7 | $60 \pm 10$ | II/III: 8/2 | $65 \pm 5$ |
| 9 Ellingsen et al. | 77 M/F:<br>63/14 | $65 \pm 22.4$ | II/III: 55/22 | $29 \pm 11.2$ |
| 10 Freyssin et al. | 12 M/F: 6/6 | $54 \pm 9$ | NR<sup>†</sup> | $27.8 \pm 4.7$ |
| 11 Fu et al. | 15 M/F: 10/5 | $67.5 \pm 1.8$ | NR<sup>†</sup> | $38.3 \pm 3.5$ |
| 12 Georgantas et al. | 20 M/F: 19/1 | $53 \pm 11$ | I/II/III: 5/14/1 | $34 \pm 11$ |
| 13 Iellamo et al. | 8 M/F: NR | 62.2 ± 8 | NR<sup>†</sup> | 31.5 ± 6.9 |
| 14 Iellamo et al. | 18 M/F: 16/2 | 67.2 ± 6 | I/II: 6/12 | 34.1 ± 6 |
| 15 Koufaki et al. | 16 M/F: 14/2 | 59.8 ± 7.4 | NR<sup>†</sup> | 41.7 ± 10.3 |
| 16 Mueller et al. | 58 M/F: 17/41 | 70 ± 7 | II/III: 44/14 | NR<sup>†</sup> |
| 17 Papathanasiou et al. | 60 M/F: 35/25 | 63.7 ± 6.7 | II/III: 48/12 | 35.9 ± 2.3 |
| 18 Roditis et al. | 11 M/F: 10/1 | 63 ± 2 | I/II/III: 3/7/1 | 30.7 ± 10.3 |
| 19 Turri-Silva et al. | 8 M/F: 5/3 | 60.9 ± 9.7 | I/II/III: 3/3/2 | 50.4 ± 17.0 |
| 20 Ulbrich et al. | 12 M/F: NR | 53.2 ± 7.0 | NR<sup>†</sup> | 28.0 ± 7.3 |
| 21 Wisloff et al. | 9 M/F: 7/2 | 74.4 ± 12 | NR<sup>†</sup> | 32.8 ± 4.8 |

\*NYHA: New York Heart Association functional class of heart failure ranging from I to □;
†NR: not reported.
‡LVFE: Left Ventricular Ejection Fraction

The exercise type selected for the HIIT protocols was primarily cycling, accounting for 14 of the studies.^21,23–27,29–32,35,36,38,39^ The next most popular types were walking uphill^33,34,40,41^ and running^22,28^, with four and two studies respectively. Only Ellingsen et al.^29^ offered a choice between running and cycling. Anagnostakou et al.^21^ included resistance strength training after their cycling protocol, whereas Papathanasiou et al.^37^ opted for high-intensity strength training combined with flexibility exercises.

According to the intensity, the most used cycling protocol is based on a peak work rate of 100% for 0.5 minutes, followed by a 0.5-minute active recovery period.^21,24,26,27,32,38^ Other protocols rate intensity for working periods at 90% Watt,^23^ 80% VO_2_max,^25,31^ 80-90% of the HRR^33,34,36^ and +15% of the first ventilatory threshold,^39^ showing a heterogeneity in the intensity measurement elected. Nonetheless, the rate of rest periods and vigorous intensity working periods for every round are in accordance with the definition of HIIT. Regarding walking uphill protocols, intensity was set from 90-95% of the peak heart rate (PHR)^40,41^ to 75-80% of the HRR^33,34^, when the running protocols matched in a 90-95% PHR.^28,29^

The studies analysed in this systematic review measure the VO_2_max using the cardiopulmonary exercise test. Other studies have measured VO_2_max to evaluate CRP effects in HF patients, as the largest multicenter, randomized controlled trial of exercise training in heart failure to date, the HF-ACTION. This study showed a moderate effect of CRP in terms of VO_2_max.^42^ Subsequently, the SMARTEX ^29^ and OptimEx ^36^ studies reaffirmed these results, although only the first one found statistical improvements of HIIT compared to moderate-intensity continuous training in this outcome. In terms of minimal clinically important difference, ten of our studies achieved a mean VO_2_max greater than 2.5 ml/kg/min.^24,26,28,30,31,33,34,37,40,41^

The total duration of the exercise per protocol ranged from a minimum of 336 minutes to a maximum of 2.160 minutes, with the longest duration being recorded. The mean duration of the complete protocols was 1.211,7 ± 430 minutes. A more exhaustive delineation of the interventions is shown in Table 2.

**Table 2.** Intervention characteristics of HIIT cardiac rehabilitation programs included in the studies.

| Study | Type | Intensity | Time/<br>session<br>(min) | Total<br>Weeks | Total<br>sessions | Total<br>time<br>(min) | VO2max Pre<br>(mL/Kg/min) | VO2max Post<br>(mL/Kg/min) |
| --- | --- | --- | --- | --- | --- | --- | --- | --- |
| 1 Anagnostakou et al. | Cycling<br>and<br>strength | alternating 0.5 min 100% pWR* with 0.5 min rest for 20 min<br>Quadriceps:3x10-12 repetitions 0.5 min rest at 55%-65% of 2RM<br>Hamstrings: 3x10-12 repetitions 0.5 min rest of quadriceps load – 1kg<br>Upper limbs:3x 10-12 repetitions 0.5 min rest at 10 RM | 40 | 12 | 36 | 1440 | 15.7 ± 4 | 17.2 ± 3.7 |
| 2 Angadi et al. | Running | 1st week: 4×2 min 80–85% PHR<sup>†</sup>; 4×2 min 50% PHR<sup>†</sup><br>2nd–4th week: 4×4 min 85–90% PHR<sup>†</sup>; 3×3 min 50% PHR<sup>†</sup> | 1st week:16<br>2nd–4th week: 28 | 4 | 12 | 336 | 19.2 ± 5.2 | 21 ± 5.2 |
| 3 Benda et al. | Cycling | 10×1 min 90% Watt<sup>‡</sup>; 10×2.5 min 30% Watt<sup>‡</sup> | 35 | 12 | 24 | 840 | 19.1 ± 4.1 | 20.4 ± 4.3 |
| 4 Besnier et al. | Cycling | 2×8 min interval training (8×0.5 min 100% PPO<sup>§</sup> alternating with 8×0.5 min 50% PPO<sup>§</sup>); 1×4 min passive recovery | 24 | 3.5 | 18 | 420 | 17.2 ± 4.5 | 20.2 ± 5.8 |
| 5 Chou et al. | Cycling | 5×3 min 80% VO<sub>2</sub> max ; 5×3 min 40% VO<sub>2</sub> max | 30 | 12 | 36 | 1080 | 15.8 ± 4 | 24.7 ± 3.2 |
| 6 Chrysohoou et al. | Cycling | 80% pWR* (pace 56-60 rpm) progressing to 100% pWR* for 0.5 min alternating with 0.5 min rest | 45 | 12 | 36 | 1620 | 16.0 ± 6.0 | 21.0 ± 5.0 |
| 7 Dimopoulos et al. | Cycling | alternating 0.5 min 100% pWR* with 0.5 min rest | 40 | 12 | 36 | 1440 | 15.4 ± 4.7 | 16.6 ± 4.9 |
| 8 Donelli et al. | Running | 4×4 min 90%–95% PHR<sup>†</sup>; 3×3 min 50–70% PHR<sup>†</sup> | 28 | 12 | 36 | 1008 | 16.1 ± 3.3 | 19.6 ± 3.5 |
| 9 Ellingsen et al. | Running<br>Cycling | 4×4 min 90%–95% PHR<sup>†</sup>; 3×3 min 50–70% PHR<sup>†</sup> | 28 | 12 | 36 | 1008 | 16.8 ± 4.4 | 18.15 ± 8.1 |
| 10 Freyssin et al. | Cycling | 1st–4th week: 3×12 repetitions of interval training (0.5 min 50% maximal power alternating with 1 min rest); 2×5 min rest<br>5th–8th week: 3×12 repetitions of interval training (0.5 min 100% maximal power alternating with 1 min rest); 2×5 min rest | 18 | 8 | 48 | 864 | 10.7 ± 2.9 | 13.6 ± 3.2 |
| 11 Fu et al. | Cycling | 5×3 min 80% VO<sub>2</sub> max ; 4×3 min 40% VO<sub>2</sub> max | 33 | 12 | 36 | 1188 | 16.0 ± 1.0 | 19.6 ± 1.2 |
| 12 Georgantas et al. | Cycling | alternating 0.5 min 100% pWR<sup>*</sup> with 0.5 min rest | 40 | 12 | 36 | 1440 | 16.6 ± 4.2 | 17.9 ± 4.7 |
| 13 Iellamo et al. | Uphill<br>walking | 4×4 min 75–80% HRR<sup>#</sup>; 4×3 min 50% HRR<sup>#</sup> | 28 | 12 | 42 | 1470 | 18.8 ± 4.6 | 23.0 ± 4.3 |
| 14 Iellamo et al. | Uphill<br>walking | 4×4 min 75–80% HRR<sup>#</sup>; 4×3 min 45–50% HRR<sup>#</sup> | 48 | 12 | 36 | 1728 | 15.3 ± 4.0 | 18.2 ± 3.0 |
| 15 Koufaki et al. | Cycling | 2×15 min cycling (1 min 20–30% PPO<sup>§</sup> alternating with 0.5 min 100% PPO<sup>§</sup>) | 30-34 | 24 | 72 | 2160-2448 | 15.3 ± 4.7 | 17.7 ± 4.9 |
| 16 Mueller et al. | Cycling | 4×4 min 80%–90% HRR<sup>#</sup>; 3×4 min active recovery | 28 | 12 | 36 | 1008 | 18.9 ± 5.4 | 20.20 ± 6.0 |
| 17 Papathanasiou et al. | Strength<br>flexibility | three high intensity intervals at 90% HRmax<sup>**</sup>; two moderate-intensity intervals at 70% HRmax<sup>**</sup> | 40 | 12 | 24 | 960 | 13.5 ± 3.8 | 17.0 ± 3.7 |
| 18 Roditis et al. | Cycling | 0.5min 100% pWR<sup>*</sup> alternating with 0.5 min rest | 40 | 12 | 36 | 1440 | 14.2 ± 3.1 | 15.4 ± 4.2 |
| 19 Turri-Silva et al. | Cycling | alternating 4 min +15% VT1 with 3 min -15% VT1*** | 28 | 12 | 36 | 1008 | 17.5 ± 4.2 | 19.6 ± 4.9 |
| 20 Ulbrich et al. | Uphill walking or running | 3-min intervals at 95% PHR interspersed by active recovery of 70% PHR | 45 | 12 | 36 | 1620 | 21.4 ± 4.1 | 24.2 ± 4.6 |
| 21 Wisloff et al. | Uphill walking | Alternating 4 min 90%-95% PHR<sup>†</sup> with 3 min 50%-70% PHR<sup>†</sup> | 38 | 12 | 36 | 1368 | 13.0 ± 1.6 | 19.0 ± 2.1 |

Exercise time calculated by the details of training protocol
\*pWR: peak work rate
†PHR: peak heart rate
‡Watt: maximal workload
§PPO: peak power output
|| VO<sub>2</sub>max: maximum oxygen consumption

### VO_2_MAX

The 21 studies selected measured VO_2_max and were included in the quantitative synthesis. The meta-analysis showed a statistically significant improvement after HIIT exercise programs (MD: 3.19, 95% CI: 2.29 to 4.09) with evidence of significant heterogeneity (p<0.01, I² = 52%). The shape of the DOI plot showed a substantial asymmetry with an LFK index of −2.04

### TOTAL MINUTES

Regarding dose-response analysis, the spline model shows a non-linear relationship between total minutes of HIIT and VO_2_max improvement (Figure 2). The model revealed a statistically significant non-linear relationship, as indicated by the chi-squared test for non-linearity (χ² = 82.44, df = 2, p < 0.001). The first spline term showed a positive association (Estimate = 0.0031, 95% CI: 0.0021 to 0.0041, p < 0.001), while the second spline term indicated a significant negative curvature (Estimate = –0.0021, 95% CI: –0.0040 to –0.0001, p = 0.038), suggesting a non-linear trend. The model was based on a single study with 21 dose levels and yielded a log-likelihood of –45.30, with AIC and BIC values of 94.59 and 96.68, respectively, which can inform model comparison. HIIT produces the greatest VO_2_max improvement at 1300 mins (MD: 3.12, 95% CI: 2.44 to 3.8)

**Figure 2.**
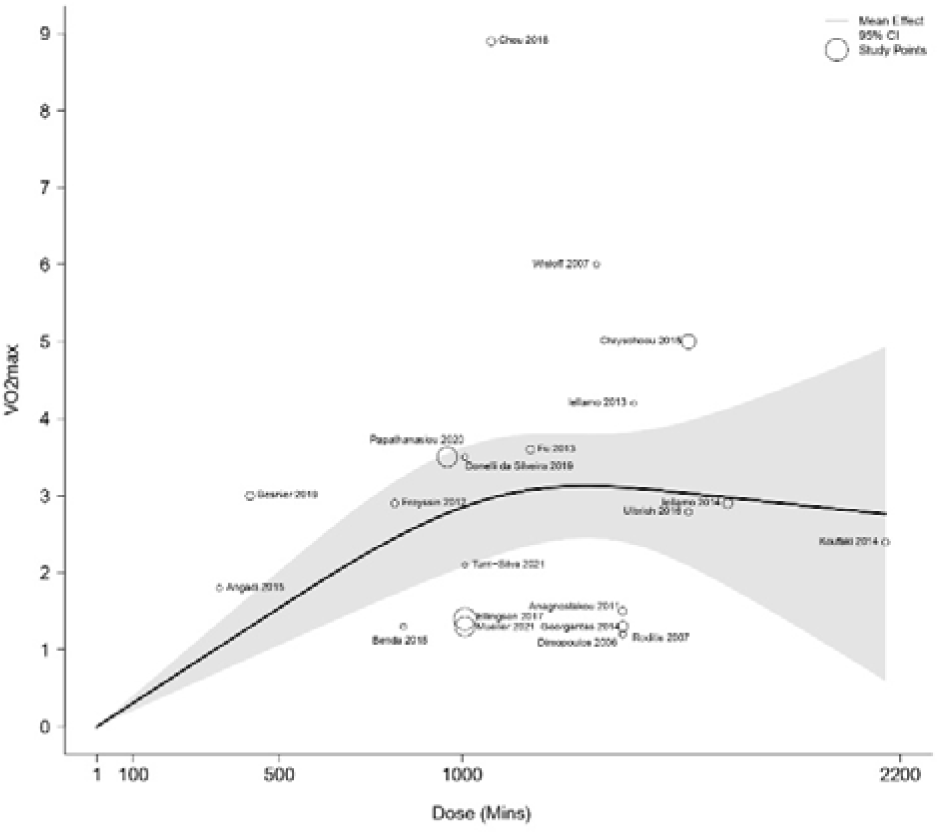
Dose-response relationship between total minutes of HIIT and improvement in VO_2_max. Studies with larger circles contributed more to the combined effect size than other studies. The solid line represents the estimate of the mean difference. The dashed lines represent the 95% confidence interval.

### MINUTES/SESSION

Regarding dose-response analysis, the spline model shows a non-linear relationship between total minutes of HIIT and VO_2_max improvement (Figure 3). Evidence of non-linearity was supported by a significant chi-squared test (χ² = 83.11, df = 2, p < 0.001). The first spline term showed a strong positive association (Estimate = 0.0949, 95% CI: 0.0643 to 0.1254, p < 0.001), whereas the second spline term, while negative in direction, was not statistically significant (Estimate = –0.0712, 95% CI: –0.1957 to 0.0534, p = 0.263), suggesting that most of the dose–response effect may be captured by the initial slope. The model was based on a single study with 21 dose levels and yielded a log-likelihood of –44.96, with AIC and BIC values of 93.92 and 96.01, respectively, to inform relative model evaluation. HIIT produces the greatest VO_2_max improvement at 50 mins per session (MD: 3.13, 95% CI: 1.35 to 4.92)

**Figure 3.**
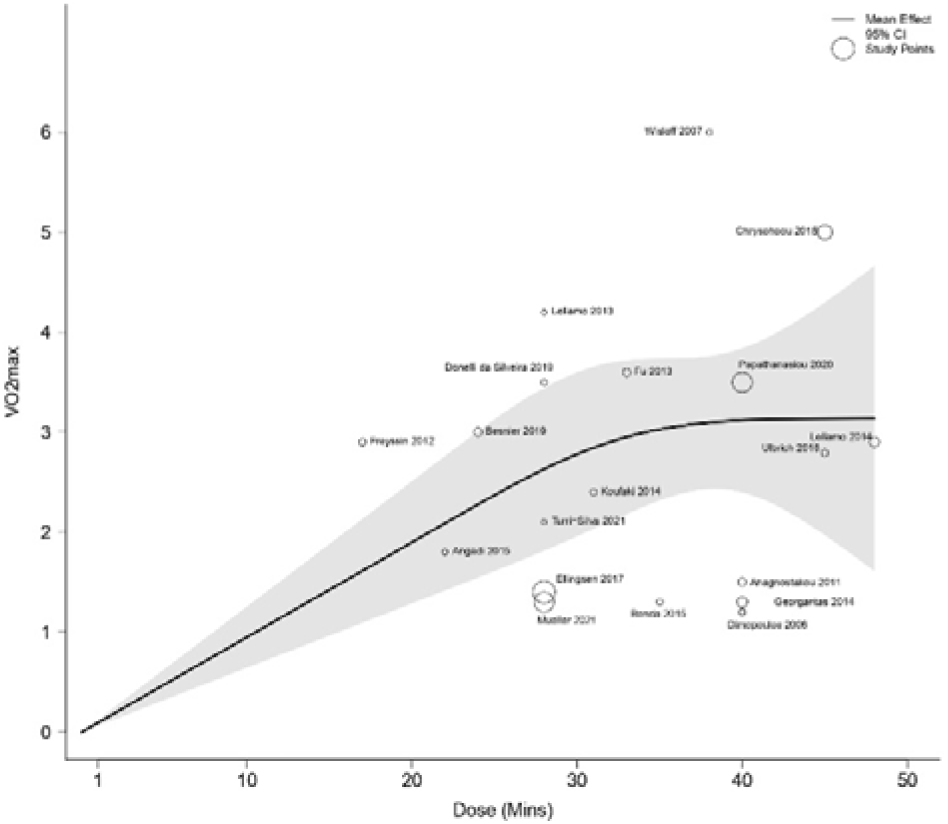
Dose-response relationship between minutes per week of HIIT and improvement in VO_2_max. Studies with larger circles contributed more to the combined effect size than other studies. The solid line represents the estimate of the mean difference. The dashed lines represent the 95% confidence interval.

### RISK OF BIAS

Overall, 28.5% of the studies had a high risk of bias, while the remaining 71.5% were rated as “some concerns”. Four studies had a high risk of bias in the missing outcome data domain (D3),^25,32,35,39^ one study had a high risk of bias in the randomization process domain (D1),^23^ and one study had a high risk of bias in measurements of the outcome domain (D4).^39^ Lastly, two studies had high risk of bias deviations from intended interventions (D2).^35,39^ The assessment of each element of risk of bias for each included study is presented in the Figure 4 and Figure 5.^43^

**Figure 4.**
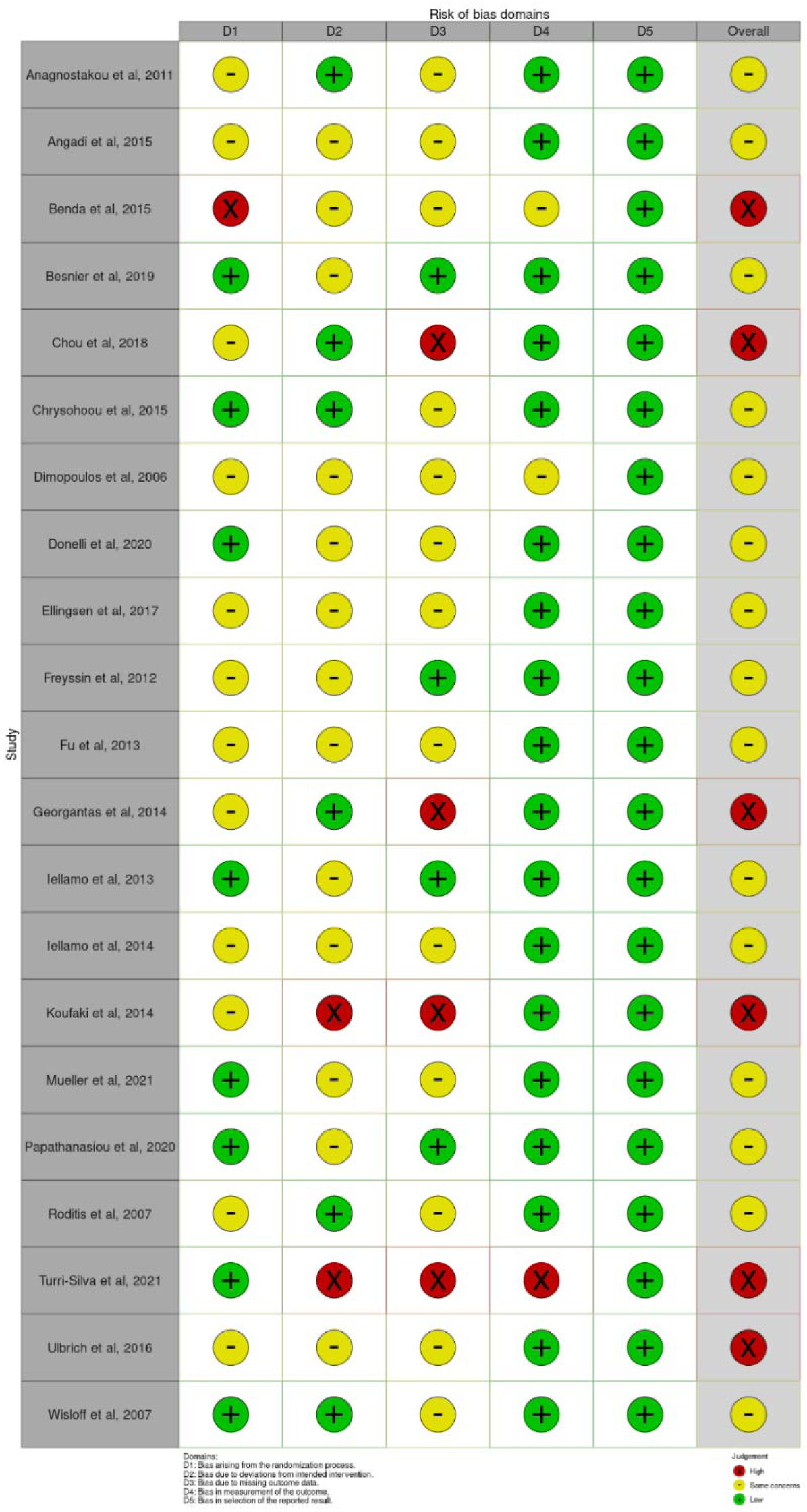
RoB2 Traffic light plot

**Figure 5.**
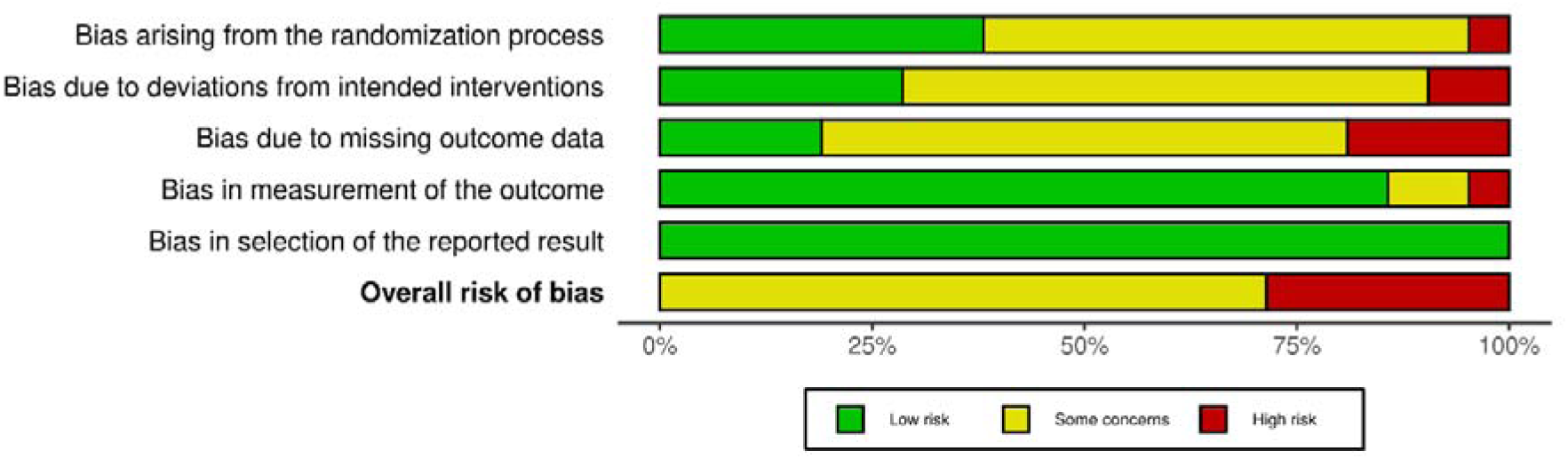
RoB2 Summary Plot

## DISCUSSION

The aim of this dose–response meta-analysis of randomized controlled trials was to evaluate the effects of the HIIT volume in CRP on VO_2_max in patients with CHF. Our main results showed not only a statistically significant improvement on VO_2_max following HIIT exercise programs but a maximum effect being achieved for this outcome after 1,300 minutes. From this point onwards, the values remain the same, with a slight decline in VO_2_max improvement up to a maximum of 2,200 minutes. A minimum dose of 30 minutes per session and a total of 900 minutes per program is necessary to overcome the minimal clinically important difference for the VO_2_max, established at 2.5 mL/kg/min,^36^, with slight increases from 35 minutes of training. However, HIIT rehabilitation programs should ideally have a duration of 50 minutes per session to optimize the VO_2_max improvements.

These findings are essential for designing effective and practical HIIT interventions. Improvements in VO_2_max have been related to reversal of concentric remodeling with subsequent improvements of myocardial contractility and compliance, as well as improved microvascular circulation and oxygen extraction.^44^ These pathoanatomical changes have been related to the improvement in clinical variables, better prognosis and higher survival rate. For example, Corra et al.,^45^ found that in stable CHF patients, 51% of subjects having a decrease in VO_2_max did not survive for the follow-up period of 1167 ± 562 days compared to 14% of subjects with an increase in VO_2_max.^46^ In addition, patients who demonstrate an increase in VO_2_max of greater than 2 mL/kg/min showed better survival rate, and each 6% increase in peak VO_2_max over three months there was a 5% lower risk for the primary outcome of all-cause mortality and all-cause hospitalization^5,47^ Therefore, HIIT interventions can directly improve patient survival and clinical outcomes, and our results could assist in optimally and time-efficient designing CRP.

Our results are consistent with proposed parameters by the ACSM, and other studies prescribing HIIT training. Unfortunately, no previous dose-response meta-analysis on HIIT volume has been conducted in CHF. Our minimum optimal recommendation of 1,300 minutes could be met with three 36-minute workouts per week over 12 weeks, ranging to 50 minutes per session to optimize VO_2_max improvements. Different reviews and meta-analysis suggest that a minimum frequency of 3 sessions per week is optimal for maximizing the benefits of HIIT regarding functional capacity.^48,49^ However, recent evidence suggests that exercise frequency is not an important determinant of cardiorespiratory fitness improvement when total weekly volume is matched.^49^ In this line, Vromen et al.^50^ found that energy expenditure appeared the only training characteristic with a significant effect on improvement in exercise capacity.^51^ Since one of the current challenges in CRP is patient adherence to CRP and it is influenced by patient preferences ^52^, it is recommended that healthcare professionals actively engage patients in the decision-making process and decide the best training frequency, considering the optimal volume recommended in this review. The adjustment of an appropriate time prescription in HIIT training, optimizing the efficiency of the time spent to obtain clinically relevant results, could be also beneficial in improving adherence to the program and lead to a better clinical result.

This systematic review and meta-analysis has some limitations. First, the patient sample from the different studies is low and not completely homogeneous, including different risk stratification stages according to the NYHA classification or even not reporting their functional class. Second, the CRP protocols were also heterogeneous in terms of FITT principles, especially in duration of the sessions and total programs. Therefore, these individual differences could influence the pooled results. Lastly, regarding the methodological assessment of the studies reviewed, the domains D2 and D3 of the RoB2 are the least met. Variability in patient compliance with the program, as well as comorbidities and complications that can result in unexpected discharges, can lead to issues with scheduling interventions and collecting data. However, VOLJmax was measured in all studies using CPET, which makes the obtained data more robust.

## CONCLUSION

This meta-analysis confirms the efficacy of HIIT in significantly improving VOLJmax in patients with CHF. The optimal dose-response was observed at 50 minutes per session and a cumulative exposure of 1300 minutes over the CRP. Notably, a minimum threshold of 30 minutes per session and 900 minutes in total was required to achieve a clinically meaningful improvement in VOLJmax. These findings underscore the significance of standardizing criteria for the design and prescription in patients with CHF, offering clinicians an evidence-based framework to optimize training prescriptions and maximize cardiorespiratory fitness.

## Data Availability

The data that support the findings of this study are available from the corresponding author upon reasonable request.

## Non-standard Abbreviations and Acronyms

CHF: Chronic heart failure
CRP: Cardiac rehabilitation programs
HIIT: High-intensity interval training
HRR: heart rate reserve
MICT: Moderate-intensity continuous training
VO_2_max: maximum oxygen consumption

